# Article Processing Charges Threaten Global Health Equity: Open Access is Closed Science

**DOI:** 10.1101/2024.11.22.24317779

**Authors:** Gabriella Y. Hyman, Taylor Wurdeman, Nikathan Kumar, Isaac G. Alty, Callum Forbes, Robert Riviello, Nakul P. Raykar

**Affiliations:** Program in Global Surgery and Social Change, Blavatnik Institute Department of Global Health and Social Medicine, Harvard Medical School; Department of Surgery, University of the Witwatersrand; Department of Surgery, Loma Linda University, Loma Linda, CA, USA; Department of Surgery, University of California San Francisco, East Bay, Oakland, CA, USA; Department of Surgery, Brigham and Women’s Hospital, Boston, MA, USA; Center for Equity in Global Surgery, University of Global Health Equity, Rwanda

**Author notes:** Corresponding author: Gabriella Y. Hyman.

## Abstract

**Introduction:** The shift from subscription-based to open-access (OA) publishing sought to democratize scientific knowledge by eliminating access barriers. However, article processing charges (APCs) have raised concerns about equity, particularly for researchers from low- and middle-income countries (LMICs). APCs shift costs from readers to authors, potentially limiting the ability of researchers to disseminate their work. This study aimed to quantify the effect of APCs on equity in global surgery by examining current APC costs and their association with journal metrics and LMIC authorship.

**Methods:** A structured review was conducted on journals’ APCs and bibliometrics using data from the Scientific Journal Rankings (SJR) and PubMed. Journals were included if they actively published human health research and had at least 100 articles with first author information. APC data were obtained from five major publishers (Elsevier, Wiley, Oxford, Sage, and Springer Nature) and supplemented manually. Journals were categorized into surgery, public health, and other groups. Data on APCs, bibliometrics (SJR ranking, H-index), and first author affiliations were analyzed using linear regression models to assess their relationship with APC costs and LMIC authorship.

**Results:** The study included 2,001 journals with a median APC of $3,700 USD showing no significant cost differences across journal categories. The median percentage of LMIC first authors was 4%, with a significant negative association between APCs and LMIC authorship: each $500 increase in APC was linked to a 0.7% decrease in LMIC first authorship. While higher APCs correlated with slightly higher bibliometric indices, the academic impact was limited.

**Conclusion:** APCs present a significant barrier to LMIC authors, undermining the equity goals of OA publishing. Alternative funding models, such as tiered pricing or expanded waivers, are needed to ensure OA remains accessible and equitable across all economic contexts.

## Introduction

The transition from subscription-based to open-access (OA) publishing sought to democratize knowledge by removing access barriers to science(1). Removing access barriers to scientific literature should promote learning, accelerate research, and advance the field by all scientists (1). However, after more than two decades of pushing towards OA, the question of whether more equity has been achieved in research is met with skepticism, often in relation to the article processing charges (APCs) (2,3). APCs, the fees publishers charge authors to publish their science in OA journals, shift the cost of disseminating literature from readers to authors (3). APCs may inadvertently restrict researchers, particularly in low- resourced settings, and low-resourced fields like global surgery (4,5).

Historically, researchers published their scholarly works without payment to journals, with journals using a subscription model or “closed access” (CA)(1). The subscription-based paradigm presented a challenge to researchers who worked for institutions that could not afford costly subscriptions to journals to access published research(BOAI 2001). This was a problem disproportionately affecting low-and-middle income country (LMIC) researchers whose institutions often did not have the resources to enter into these subscription agreements(1). With the advent of technological advancements, scientists and journals supported the move towards free and unrestricted online availability of information. This led to the mainstreaming of OA models with journals offering OA, subscription-based, or hybrid options (publication beginning as subscription-based for a limited embargo period before moving to OA)(1,6).

The OA model exists within the Creative Commons license rules governing the dissemination of scientific material (7). Additionally, academic and funding institutions often make OA a requirement for publishing. APCs are intended to support the financial sustainability of OA journals and add value for researchers (such as faster processing time and broader dissemination of findings), yet they introduce a financial burden that may affect equity in scientific research (3,8). While APCs make research available to authors without institutional affiliation for viewing, they may be prohibitive for the same researchers, particularly those in LMICs who wish to publish their scientific work(9). Considering less than 10% of global investment in health research is spent in LMICs, the burden of payment for publishing research OA disproportionately affects researchers in LMICs where income is also significantly lower (10,11) .

Amid growing concerns among the academic global surgery community that APCs are becoming increasingly unaffordable, evidence to improve understanding of the current APC model is needed. In global health fields, such as academic global surgery, equity is a guiding principle and so instruments that challenge equity are problematic. In addition to the cost of publishing, researchers also consider the potential impact of a journal when selecting a journal. Journal bibliometrics, such as Scientific Journal Rankings (SJR), impact factor (IF), or H-index, are used as proxies for quantifying the value of publishing with a specific journal(12). This study aims to quantify current APC costs and assess the impact of APCs on equity and journal bibliometrics in global surgery.

## Methods

### Overview

We performed a structured review on journals’ APCs and bibliometrics. We extracted journal ranking from the Scientific Journal Rankings (SJR) database and cross-referenced these with journals indexed in PubMed (13). Journals were included if they were present in both databases, were actively publishing (meaning the most recent article published was on or after 1 February 2023), and if they had at least 100 articles with first author information listed. Journals were excluded if the SJR category did not pertain to “human health”. Journals were categorized as open access (OA), hybrid, or closed access (subscription-based). No journals uniformly defined the definition of “hybrid”. Data on the following bibliometrics were available and included: SJR ranking and H-index. SJR is the average number of weighted citations received in a given year against the citations for documents published in that journal over the previous 3 years (14). A journal’s H-index is the ratio of a journal’s total articles (*h*) which have received at least *h* citations(15).

We identified five major publishing houses that provided open-sourced datasets of APCs at the journal level (Elsevier, Wiley, Oxford, Sage and Springer Nature) and manually retrieved them for JAMA. These datasets were matched to the SJR dataset. Journals were categorized based on their SJR category into three groups: surgery, public health, and other. A journal had one category assigned for analysis, and a surgery categorization was prioritized over public health or other due to the primary focus of this study being on global surgery. Where not available in publishing houses datasets, information on the publishing model was retrieved through purposive searching by authors GH and TW.

### Data handling

RStudio version 4.4 was used for analysis. Using the *retrez* package, we extracted article metadata from the 200 most recent articles for each included journal. This included first author affiliation, which was analyzed for country of affiliation. Country affiliation was categorized by income level according to the World Bank Group’s classifications as LMIC (which included low-income, low-and middle-income, and upper-middle- income countries) or high-income country (HIC)^10^. Each journal’s *LMIC First Author percentage (LMIC %),* which was used as a proxy for equity, was calculated by dividing the number of articles with an LMIC first author by the total number of articles. Articles were excluded from the denominator if the first author did not have an affiliation listed.

The final dataset contains the following variables: SJR ranking, H-index, country of the journal, country of the publisher, SJR journal category, first author affiliation percentage (*LMIC %*), publishing model (OA or hybrid), APC ($USD), and creative common (CC) license.

### Data analysis

For descriptive analysis, we depicted continuous data using median and interquartile range, and for categorical data we used percentage and frequencies. Descriptive statistics are presented for the overall dataset and for each defined journal category: surgery, public health, and other.

We used a linear regression to describe the relationship between APC and bibliometrics (SJR and H-index). Both bibliometric values were included as validity measures for the study. We developed models to analyze the relationship between standardized bibliometric rank and LMIC first authorship percentage. We included standardized APC, SJR, and open access status as covariates. An interaction term was used between the standardized bibliometric rank and standardized APC, as well as for access model (OA versus hybrid). We standardized SJR ranking and H-index by subtracting the overall mean from each value and dividing by the standard deviation. We standardized the APC by dividing it by $500. This allows for an increased ease of interpretation of the model, whereby we provide a unit of measurement ($500) for APCs for each unit increase or decrease in the model coefficient.

In order to demonstrate the relationship between median APC cost compared to income of LMIC medical professionals, we constructed a color-coded map using APC as a percent of gross national income (GNI) per capita (16).

## Results

### Overview

The final dataset consisted of 2001 journals (**Figure 1**). The full list of journals can be found in **Appendix 1**. The majority of journals were in the *other medicine* category (n=1552), with 9% (n=180) in the *public health* category and 13% (n=269) in the *surgery* category. No journal reported an exclusively subscription-based publication model, with 83% (n= 1658) reporting a hybrid over a pure OA model. The overall median APC was $3700 (IQR= $1030) with no significant difference between journal categories (p=0.2). The median SJR and H-index were 0.87 and 89 respectively (**Table 1**)

**Figure 1:**
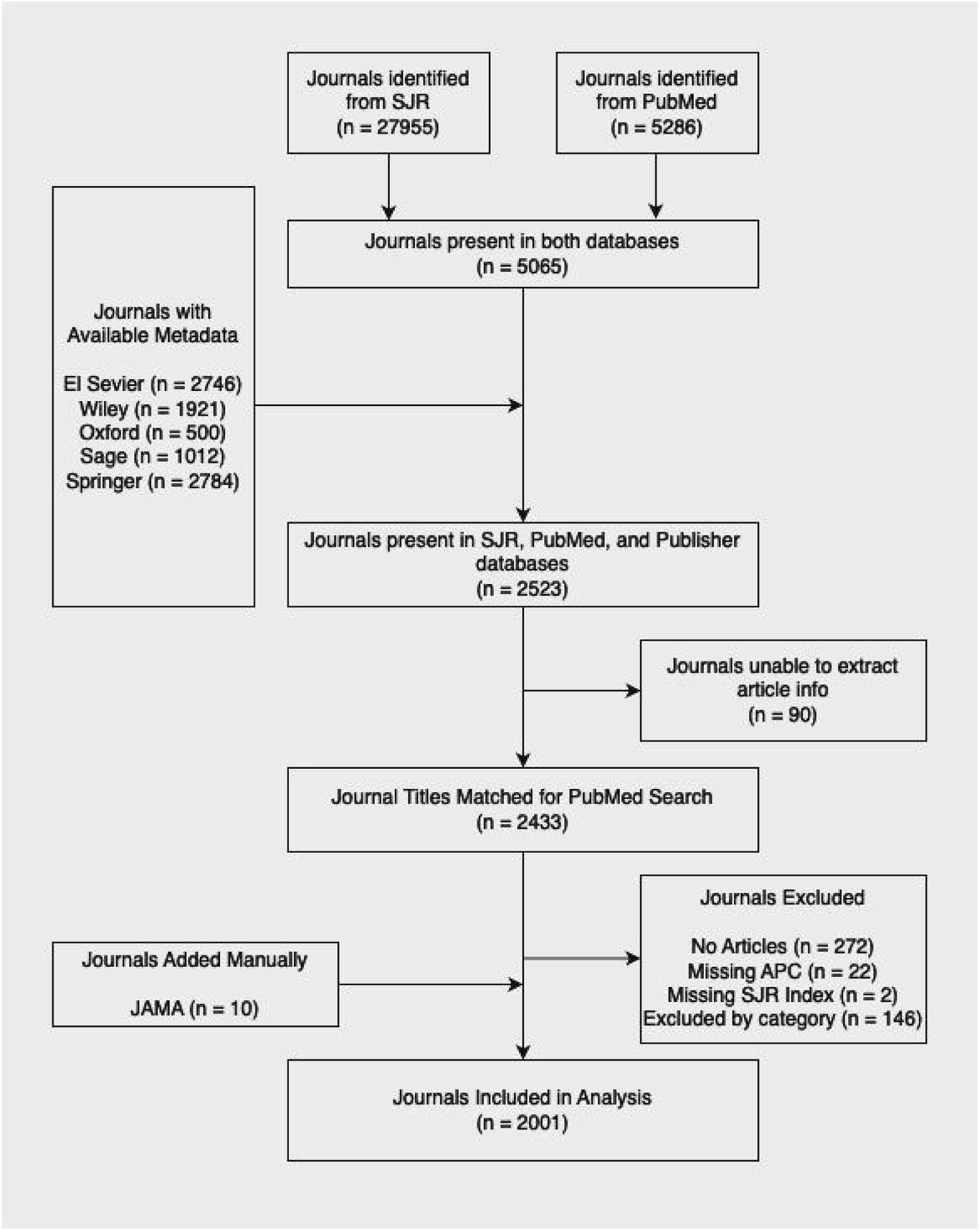
Study flow diagram showing journals included in the analysis.

**Table 1:**
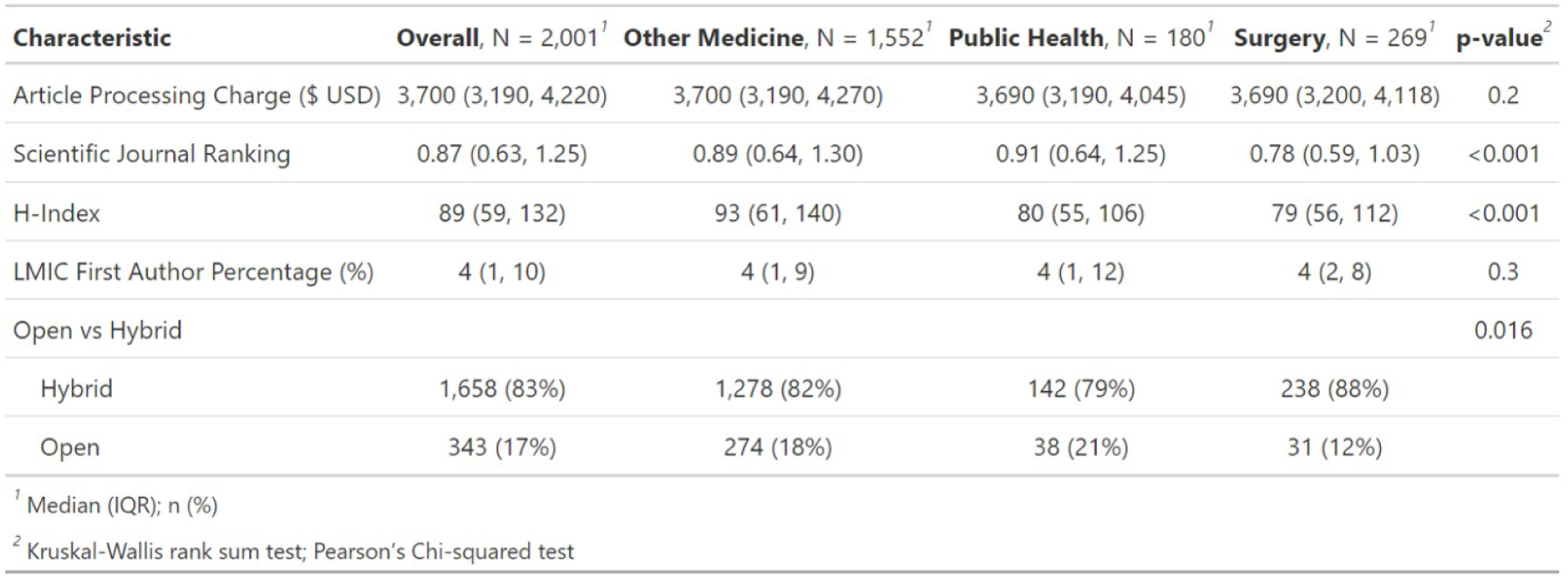
Characteristics of Included Journals.

| Characteristic | Overall, N = 2,001<sup>1</sup> | Other Medicine, N = 1,552<sup>1</sup> | Public Health, N = 180<sup>1</sup> | Surgery, N = 269<sup>1</sup> | p-value<sup>2</sup> |
| --- | --- | --- | --- | --- | --- |
| Article Processing Charge (\$ USD) | 3,700 (3,190, 4,220) | 3,700 (3,190, 4,270) | 3,690 (3,190, 4,045) | 3,690 (3,200, 4,118) | 0.2 |
| Scientific Journal Ranking | 0.87 (0.63, 1.25) | 0.89 (0.64, 1.30) | 0.91 (0.64, 1.25) | 0.78 (0.59, 1.03) | <0.001 |
| H-Index | 89 (59, 132) | 93 (61, 140) | 80 (55, 106) | 79 (56, 112) | <0.001 |
| LMIC First Author Percentage (%) | 4 (1, 10) | 4 (1, 9) | 4 (1, 12) | 4 (2, 8) | 0.3 |
| Open vs Hybrid |  |  |  |  | 0.016 |
| Hybrid | 1,658 (83%) | 1,278 (82%) | 142 (79%) | 238 (88%) |  |
| Open | 343 (17%) | 274 (18%) | 38 (21%) | 31 (12%) |  |

<sup>1</sup> Median (IQR); n (%)
<sup>2</sup> Kruskal-Wallis rank sum test; Pearson's Chi-squared test

### Cost and Equity

The overall median LMIC First Author Percentage was 4% (IQR= 10), with no statistically significant difference between journal categories (p=0.2) (Table 1). Of the 203 countries with data available, APCs constituted less than 5% of GNI per capita in only 7 countries. In 63 countries, the median APC constituted >95% of GNI per capita (Figure 2).

**Figure 2:**
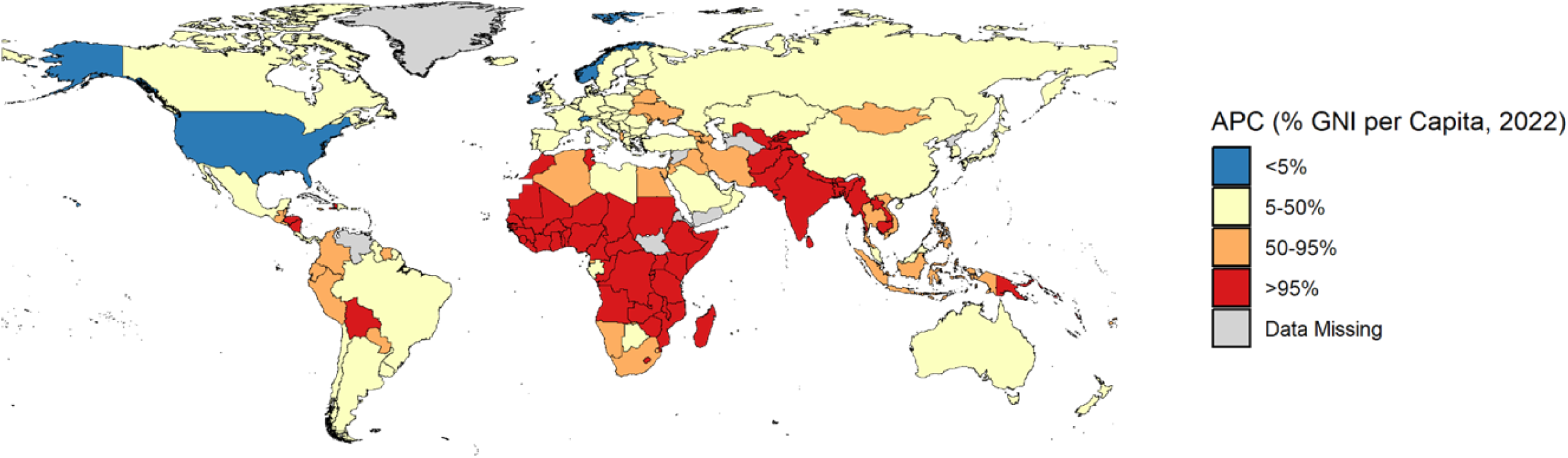
Mean APC as a percentage of GNI per Capita.

One standard deviation increase in SJR was associated with a 1.14 unit increase in APC, where each unit represents $500 (p<0.01). One standard deviation increase in H-index was associated with a 1.16 unit increase in APC (p<0.01) .

Table 2 depicts the models with LMIC first authorship percentage as a dependent variable. In model 1, a one-standard deviation increase in SJR was associated with a 2.7 percentage point (67.5%) decrease in LMIC authorship (p < 0.01). An increase in APC by $500 was associated with a 0.7 percentage point (17.5%) decrease, in LMIC authorship. Surgery journals were associated with a drop in LMIC authorship percentage (1.4 percentage points, p < 0.05) (Figure 3). The publication model (hybrid versus open) was not associated with LMIC authorship percentage (p = 0.69). The positive coefficient of the interaction term between SJR and APC indicates that the negative association of APC on LMIC authorship diminished at higher levels of SJR. Model 2, which substitutes H-Index for SJR, demonstrated similar results for all covariates, with small differences in coefficients (Table 2, Figure 4).

**Figure 3:**
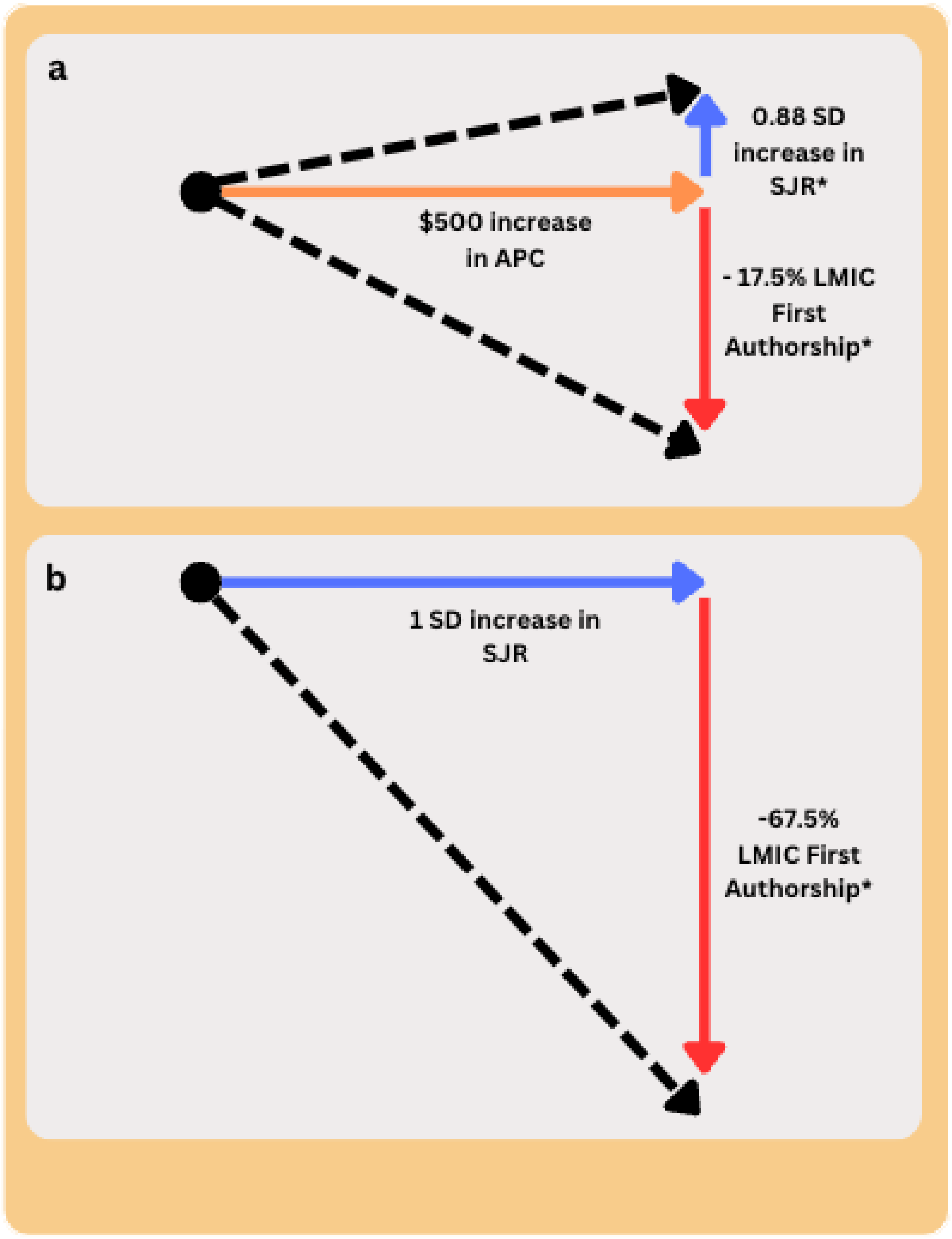
Association between APCs, journal impact and LMIC first authorship.

**Figure 4:**
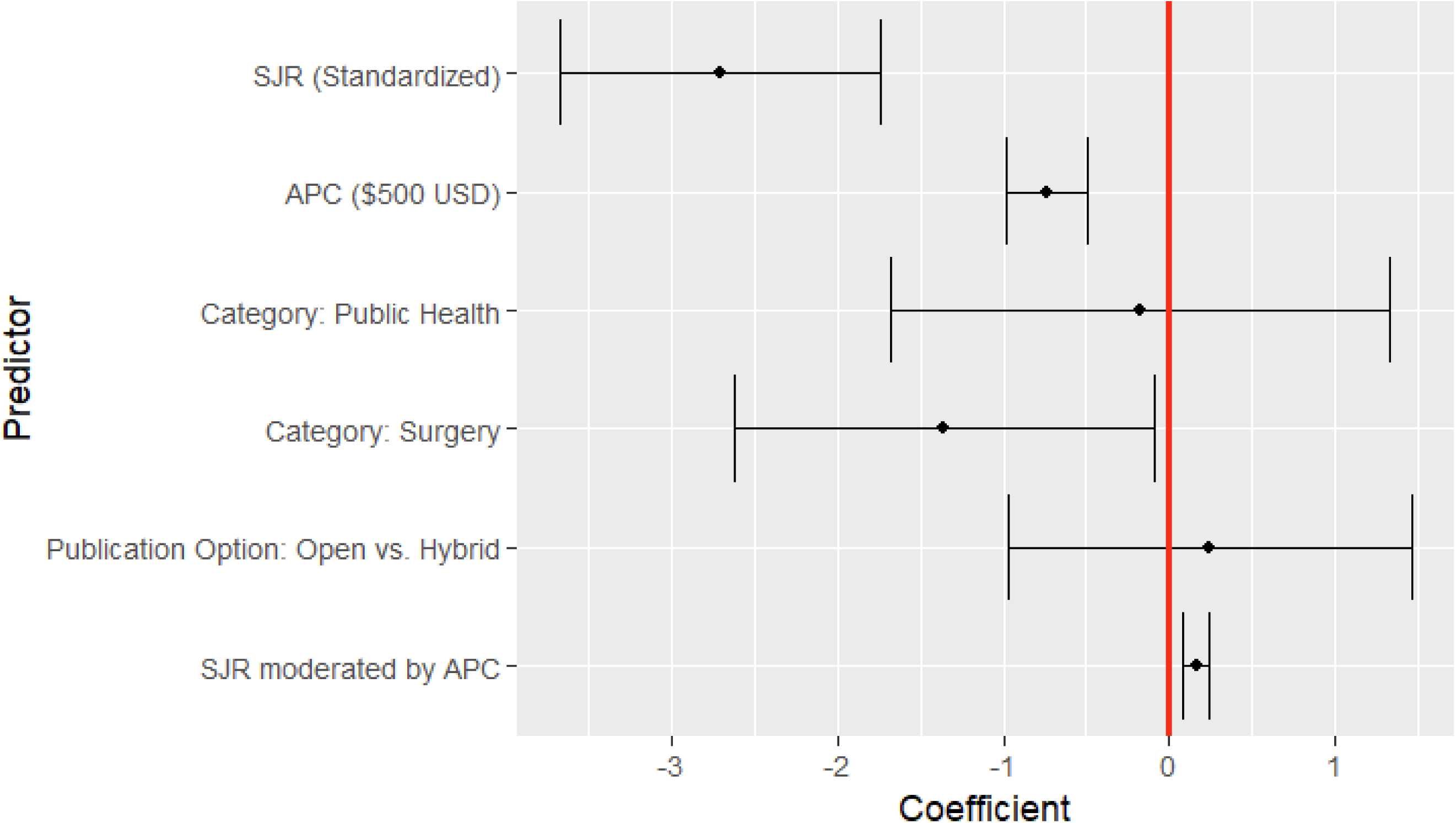
Effect of increasing SJR and APCs on LMIC first authorship.

**Table 2:**
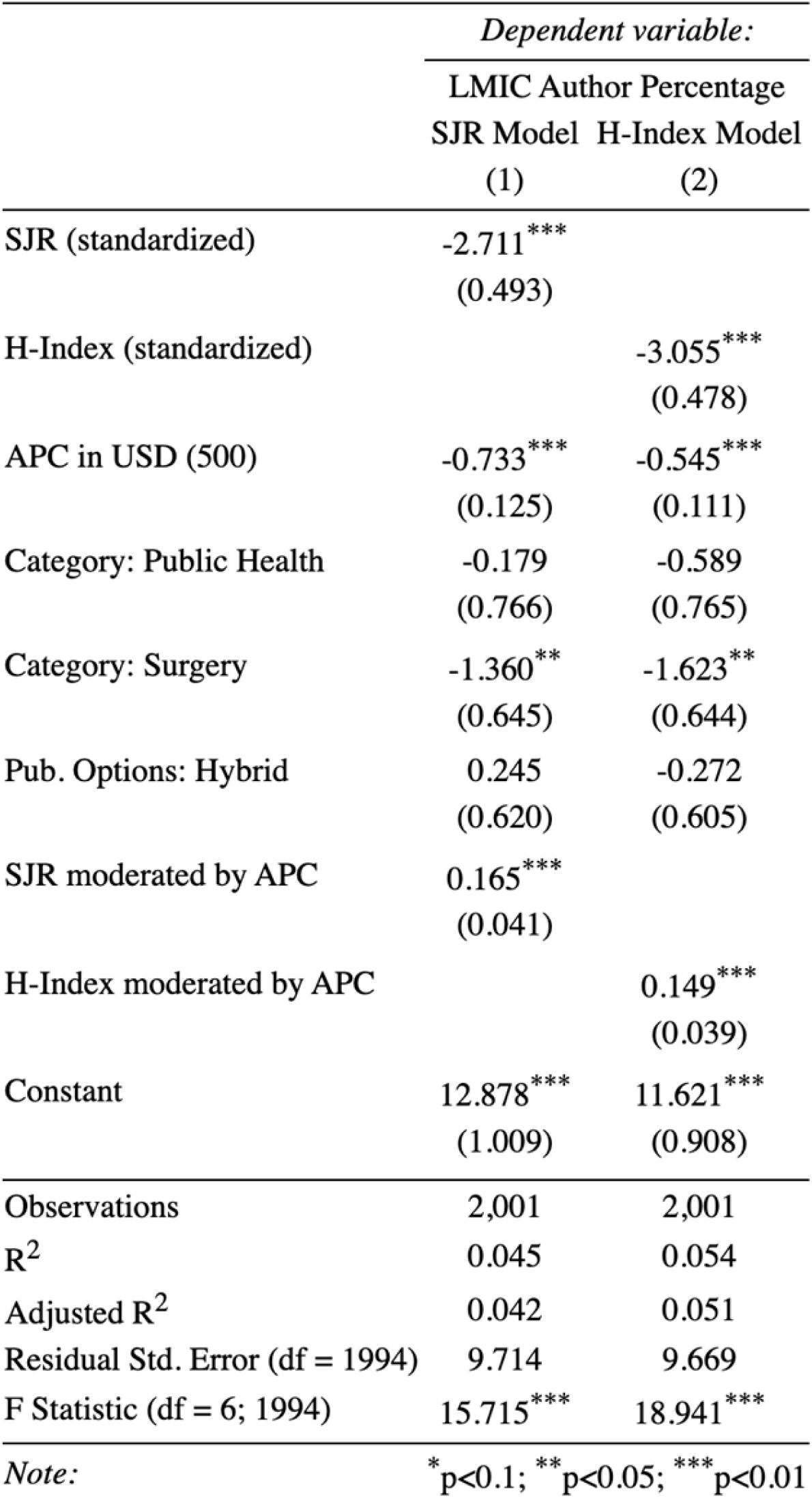
Model predicting the effect of SJR and H-index on LMIC First Authorship Percentage.

### Publishing Options

Several journals reported multiple creative commons (CC) licensing options for researchers (Table 3). Of the 1094 for which CC licensing was retrievable, most offered the option with the most researcher rights. Some journals offered more than one CC option. CC licensing options provide a standardized way to describe user and owner rights of intellectual property. This guides users about what they can do to use, disseminate and shared published work (7,17).

**Table 3:**
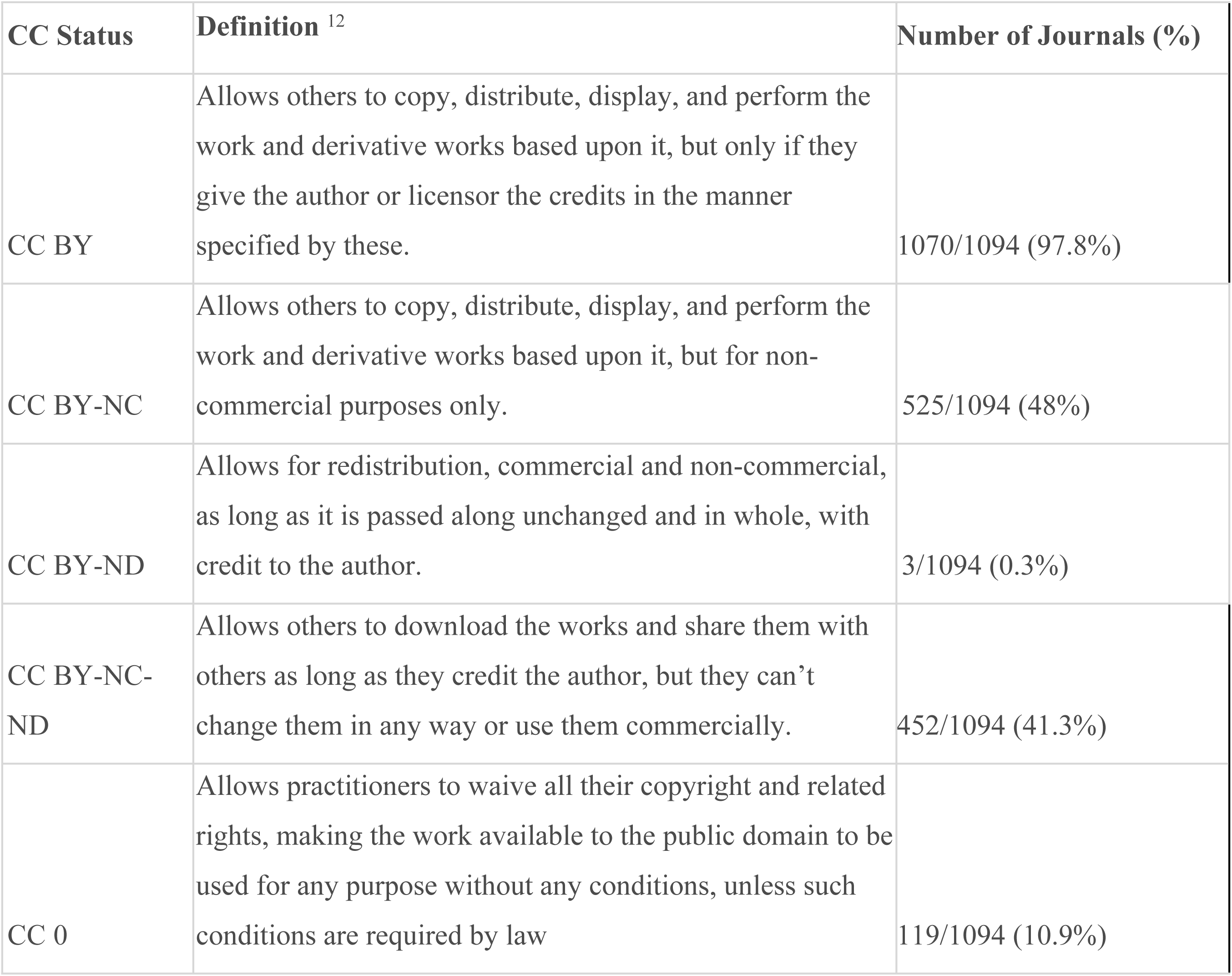
Distribution of Creative Commons (CC) License options among Journals.

Waiver options were dependent on the publishing company for each journal. The five different waiver options were variable and dependent on country or institutional agreements, or the proportion of authors from LMICs. The use of waiver options is not reported with each specific journal article thus we were not able to evaluate the effect of waiver options on publication practices.

Journals were either categorized as fully open access or hybrid. Fully open access journals do not offer traditional subscription-based options, and instead require either no APC, an institutional agreement for APC waivers, an APC waiver based on author criteria, or payment of the full APC for publication. Hybrid journals offered both open access and traditional subscription-based models. The subscription-based model option typically comes with no APC but has access to the public restricted, or delayed by months or years.

## Discussion

### Overview

This structured review describes the implications of Article Processing Charges (APCs) on researchers in the field of Global Surgery. The stated goal of OA publishing, which uses APCs as a financing method, is to improve accessibility of scientific research, and to make the scientific and publishing processes more transparent and inclusive (1,18). Our findings regarding APC costs and the subsequent equity implications are described in three key messages: (1) the financial burden of APCs is high; (2) APCs have a negative impact on authorship equity, particularly affecting first authors from LMICs; and (3) the correlation between bibliometrics and APCs is of limited practical, academic significance.

### Financial Burden

Our findings suggest that APCs are high. When compared to countries’ GNI per capita, we found that only a small percentage of researchers (in the USA and Scandinavian countries) publish research where APCs constitute less than 5% of GNI per capita. This means that where APCs are incurred as out-of-pocket expenses for researchers, their high cost can be prohibitive for researchers who may have to prioritize which data to publish, or even whether to submit for publication at all.

Alternatives to what is perceived as the conventional APC model exist. Authors have various dissemination options in accordance with CC restrictions and waiver options(7,17). However, journals do not report whether a publication was published using a waiver option or not, making the objective evaluation of whether authors use waiver or self-cite options difficult. Furthermore, there is often no guarantee of a waiver at the start of the submission process. Thus, high APCs may deter researchers from submitting their research to a particular journal altogether. While we were unable to evaluate the disparate definitions of the “hybrid” designation given by journals and publishers, it is possible that even when only certain article types are not subject to mandatory APC charges for OA publishing, or that only certain article types have the option of being published behind a paywall, a journal is designated as ‘hybrid’. We conducted further ad-hoc searches which revealed that many journals offer hybrid options only for certain article types, maintaining an OA publishing model requiring APCs for a great portion of their offerings.

Thus, the designation of hybrid may overestimate the true options available to researchers within the current OA model.

### Equity Implications

Our analysis suggests the current APC model does not achieve its stated intentions of achieving research equity. Despite the initial promise of OA to democratize science for all, our findings suggest that science has been commoditized by the high cost of APCs (1). APCs are high across different journal categories and publishing houses and perpetuate inequities in academia that disproportionately affect researchers from LMICs (5,19). Compared with a review conducted by Morrison et al in 2021, which found the average APCs across all disciplines to be $958 and for the field of medicine to be $1373, our study shows a much higher median amount of $3700, even for global health journals (Morrison et al. 2022). While our methodology was different in terms of our search, journal categorization and focus on specific publication houses, all of these APC amounts exist in stark contrast to the field of global health’s core principle of equity and present a barrier to advancing scientific advancements.

An increase in APC by $500 is associated with a 0.7 percentage point decrease in LMIC first authorship. Given the average LMIC authorship is 4%, these values represent large relative changes in LMIC authorship percentages of 17.5% (Figure 3). This is more pronounced for specialty surgery journals where LMIC first authorship dropped by 35%. This raises concerns about the exclusionary effects of APCs. A previous review discussed a number of potential barriers to achieving publication equity (19). Possible reasons for LMIC first authorship percentage being negatively affected by APCs are that LMIC authors are more sensitive to changes in price ($500 unit increases) as APCs are a high proportion of their annual salary. Given that APCs are often paid out of pocket and much of the global surgery research in the global south is unfunded, the expectation to pay APCs out of pocket would almost certainly preclude LMIC first authors from the OA APC-funded model (4,10,19). It is worth noting that LMIC first authorship is affected despite waiver options described for LMIC researchers (Table 3) (5) .

Several institutions and funders of biomedical research have taken steps towards making APCs more affordable for researchers (21–26). However, these funding agreements between biomedical funders and publishing houses only support employees or researchers funded by these institutions. This leads to an increase in the proportion of researchers publishing research which has institutional funding support.

Many LMIC researchers are not funded through grant support, and within HICs, only a small proportion of grant-funding is for global health research(11,27). Moreover, an additional layer of inequity is introduced into funding cycles as researchers with large grants or institutional support are more likely to be able to afford APCs, leading to more publications, and further funding.

### Academic Significance

Bibliometrics are the quantitative analysis of academic publications to evaluate the publication in terms of number of citations and ‘impact’ (28). SJR measures the scientific influence of the average article in the journal by calculating the weighted number of citations in a given year to citable publications published in the journal within the three preceding years (14) . H-index measures both the productivity and citation impact of the publications of a scientist (15). The underlying assumption is that higher values in these metrics reflect a greater research ‘impact’, suggesting that the work has been frequently referenced and deemed influential by the academic community. This is often a core goal of researchers when selecting a publication route. We found the relationship between APC and both bibliometric measures shows a weak positive association for both H-index and SJR. The idea that this incremental increase is of academic significance is not convincing relative to the steep associated increase in APC cost (Figure 3).

While OA was designed to resolve inequities in a subscription-based model through making research more accessible for all readers, the current OA model has shifted the financial burden of publishing to the author. This shift, which has led to higher APCs, has a significant financial impact on authors from LMICs, and is associated with lower rates of LMIC first authorship. Therefore, the current OA model has shifted the financial burden to a group that also has limited resources - LMIC authors. The financial burden of APCs is not negligible in high-resourced settings either. For example, in Germany and the United Kingdom, which are among the top producers of scientific publications according to the National Science Foundation, APCs make up between 5-50% of GNI per capita (29). This means that APCs can be prohibitive for researchers, who may be forced to choose between paying a salary, conducting further research, or paying for APCs. The advancement of scientific enquiry, whether intended to improve clinical outcomes or strengthen health systems, benefits from a broad range of diverse perspectives. The effects of APCs on decreasing authorship diversity based on financial resources, therefore weakens science in addition to being inequitable.

### Implications and Future Directions

There are several options for researchers to be both cost-saving and impact-driven within the current OA system. While authors may experience a sense of pressure to pay APCs, authors should reconsider the options for circumventing APC costs because the OA model is not achieving its stated goals of accessibility and inclusivity. APCs are not furthering equity, as shown by our LMIC first authorship proxy, nor is there strong evidence that it is helping advance science in a way that is academically significant as shown by our bibliometric proxy. With growing pressure from large biomedical research agencies in the European Union and the United States (NIH) to publish OA, there is an opportunity to call for an increase in transparency and equity(25). Authors have various choices to publish within the diverse options laid out by the CC-BY framework(17,20,24). Several journals provide multiple licensing pathways that would allow for the dissemination of scientific work outside the traditional OA publishing model, such as self-archiving. Self-archiving can take place prior to or after publication depending on the journal and access type (OA or subscription-based) and allows authors to share pre-print versions of their research prior to peer-review and publication (30,31). More education and community-sensitization is needed to improve authors’ knowledge of the CC options available to them and to improve our understanding of the factors affecting author’s decisions to publish OA.

### Limitations

Our study has several limitations. Firstly, journals analyzed are predominantly managed by publishing houses in HICs, which may reflect a bias towards HIC perspectives and priorities. Factors such as advertising, journal visibility, and author factors may affect citations more than the OA model itself and may affect researchers’ journal choice. The low number of LMIC first author’s overall is linked to complex factors extending beyond only APCs, requiring further evaluation. Waiver options for publishing houses were included as opposed to individual journals. Further research is needed to improve understanding around the complex factors that affect author choices of journals.

## Conclusion

This review highlights financial and equity challenges posed by APCs in global surgery research. We found that APCs impose a high financial burden and negatively affect authorship equity, particularly for researchers from LMICs, with little justification provided by the weak correlation between bibliometrics and APCs. These findings call for the exploration of alternative models such as tiered pricing and more widespread availability of waivers to ensure that open access fulfills its promise of democratizing knowledge across all economic contexts. This approach is essential for fostering a truly equitable academic publishing environment.

## Data Availability

All primary data was obtained from open-source databases: Scimago Journal and Country Rank (SJR) Database of indexed journals bibliometrics (SJR and H-index) PubMed and National Library of Medicine complete list of indexed journals. APC data were obtained from major publishers’ websites’ primary mega-data and supplemented manually for: Elsevier, Springer-Nature, JAMA, Oxford University Press, Sage, Taylor-Francis: Peeref. “Top 10 Largest Academic Publishers in 2022.” https://www.peeref.com/collections/top-10-largest-academic-publishers-in-2022. Elsevier. “Policies and Standards – Pricing.” https://www.elsevier.com/about/policies-and-standards/pricing#3-no-double-dipping. SAGE. “SAGE Choice - Open Access Options.” https://us.sagepub.com/en-us/nam/sage-choice. Springer Nature. “Open Research: Journals and Books.” https://www.springernature.com/gp/open-research/journals-books/journals. Oxford University Press (OUP). “Charges, Licences, and Self-Archiving.” https://academic.oup.com/pages/open-research/open-access/charges-licences-and-self-archiving. Taylor & Francis. “Open Access Pricing.” https://taylorandfrancis.com/our-policies/open-access-pricing/. Taylor & Francis. “Open Access Cost Finder.” https://authorservices.taylorandfrancis.com/choose-open/publishing-open-access/open-access-cost-finder/?utm_source=CPB&utm_medium=cms&utm_campaign=JPJ17055&_ga=2.139322611.1368322867.1710764306-1168379644.1710764306&_gl=1zp6flq_gaMTE2ODM3OTY0NC4xNzEwNzY0MzA2_ga_0HYE8YG0M6MTcxMDc2NDMwNi4xLjAuMTcxMDc2NDMwNi4wLjAuMA.._gcl_au*NzUzOTYwNjc5LjE3MTA3NjQzMDY. Wiley. “Article Publication Charges.” https://authorservices.wiley.com/author-resources/Journal-Authors/open-access/article-publication-charges.html. Publishing houses creative commons licenses: Peeref. “Top 10 Largest Academic Publishers in 2022.” https://www.peeref.com/collections/top-10-largest-academic-publishers-in-2022. Elsevier. “Policies and Standards – Pricing.” https://www.elsevier.com/about/policies-and-standards/pricing#3-no-double-dipping. SAGE. “SAGE Choice - Open Access Options.” https://us.sagepub.com/en-us/nam/sage-choice. Springer Nature. “Open Research: Journals and Books.” https://www.springernature.com/gp/open-research/journals-books/journals. Oxford University Press (OUP). “Charges, Licences, and Self-Archiving.” https://academic.oup.com/pages/open-research/open-access/charges-licences-and-self-archiving. Taylor & Francis. “Open Access Pricing.” https://taylorandfrancis.com/our-policies/open-access-pricing/. Taylor & Francis. “Open Access Cost Finder.” https://authorservices.taylorandfrancis.com/choose-open/publishing-open-access/open-access-cost-finder/?utm_source=CPB&utm_medium=cms&utm_campaign=JPJ17055&_ga=2.139322611.1368322867.1710764306-1168379644.1710764306&_gl=1zp6flq_gaMTE2ODM3OTY0NC4xNzEwNzY0MzA2_ga_0HYE8YG0M6MTcxMDc2NDMwNi4xLjAuMTcxMDc2NDMwNi4wLjAuMA.._gcl_au*NzUzOTYwNjc5LjE3MTA3NjQzMDY. Wiley. “Article Publication Charges.” https://authorservices.wiley.com/author-resources/Journal-Authors/open-access/article-publication-charges.html. World Bank Group data: GNI per capita for all countries

## Disclosures

None

## Funding

Nil

## Contributions

Conceptualization and design of the study: GH, TW, CF, NR

Literature review and synthesis: GH, TW

Initial data collection and management: GH, TW

Methodology design: GH, TW

Quantitative statistical analysis: GH, TW

Final data cleaning, review and interpretation: GH, TW, NK, IA, CF, NR

Manuscript writing and revision: GH, TW, NK, IA, NR

Review and editing of the manuscript: GH, CF, RR NR, NK, IA

Final approval and revision: GH, RR, NR

## Notes

### Competing Interest Statement

The authors have declared no competing interest.

### Funding Statement

The author(s) received no specific funding for this work.

### Author Declarations

Secondary data analysis of open source data. No IRB application made.

